# Genetic Evaluation in Chronic Kidney Disease: A Canadian Single-Centre Experience from a Multicultural Urban Population

**DOI:** 10.1101/2025.11.07.25339114

**Authors:** Zachary T. Sentell, Felicia Russo, Marc Henein, Lina Mougharbel, Zachary W. Nurcombe, Ahsan Alam, Dana Baran, Lorraine E. Bell, Daniel Blum, Marcelo Cantarovich, Andrey V. Cybulsky, Sonali De Chickera, Mallory L. Downie, Bethany J. Foster, Gershon Frisch, Paul R. Goodyer, Indra R. Gupta, Laura Horowitz, Serge Lemay, Mark L. Lipman, Sharon J. Nessim, Tiina Podymow, Ratna Samanta, Shaifali Sandal, Rita Suri, Tomoko Takano, Emilie Trinh, Murray Vasilevsky, Elena Torban, Ruth Sapir-Pichhadze, Thomas M. Kitzler

## Abstract

**Background:** Chronic kidney disease (CKD) affects over 10% of the global population. A genetic diagnosis can be identified in about 30% of pediatric and 10-30% of adults, informing treatment, prognosis, and family-based risk assessment. However, access to renal genetics services remains limited across many healthcare systems.

**Objectives:** To characterize the clinical and genetic landscape of CKD in patients referred for genetic evaluation within a Canadian single-centre nephrology-genetics program, and to evaluate the diagnostic yield and clinical utility of an integrated renal genetics clinic.

**Methods:** We conducted a retrospective study of 206 probands referred for suspected hereditary kidney disease to the McGill University Health Centre Renal Genetics Clinic between 2019 and 2024. Genetic testing was performed in accredited laboratories, predominantly through comprehensive multi-gene panels or phenotype-directed exome sequencing. All reported variants were classified according to the ACMG/AMP criteria, and variants of uncertain significance were reevaluated *post hoc* using standardized quantitative and evidence-tier frameworks to determine whether they trended toward “likely pathogenic” (“hot”) or “likely benign” (“cold”), without implying formal reclassification.

**Results:** A molecular diagnosis was established in 34.5% of probands (71/206), implicating pathogenic or likely pathogenic variants across 35 genes representing diverse monogenic kidney disease etiologies. The highest diagnostic yields were observed in cystic nephropathy (51.9%), tubulopathy (38.5%), and glomerulopathy (35.6%). Genetic results affected clinical management in 23.9% of diagnosed cases, leading to changes in treatment for 16.9%, modification of transplant management in 5.6%, informed living donor risk assessment in 14.1%, and facilitated cascade testing in 66.2% of families. CKD of unknown etiology comprised 28% of the cohort, with a genetic diagnosis reached in 25.9% of these cases. Variants of uncertain significance (VUS) were reported in 39.3% of probands, with higher overall variant burden and lower diagnostic yields among individuals of non-European ancestry. *Post hoc* internal reassessment stratified 67.7% of VUS as mid or lower confidence (“cold”) and 32.2% as higher confidence (“hot”) or likely pathogenic.

**Conclusions:** In a diverse urban population, integration of a dedicated renal genetics service within nephrology care achieved high diagnostic yield, substantially influenced management, and facilitated family risk assessment. Structured referral pathways and multidisciplinary variant interpretation optimize the clinical utility of genetic testing in CKD.

## Introduction

Chronic kidney disease (CKD) affects about 850 million people globally, and approximately 1 in 10 Canadians (roughly 4 million people), causing significant morbidity and mortality, while exerting a tremendous toll on the health care system.^1–5^ Genetic testing in patients with suspected inherited kidney disease can transform clinical care by providing definitive molecular diagnoses that inform personalized treatment decisions, family counseling, and prognostic assessment.^6–10^ Monogenic disorders underlie about 10-20% of adult CKD and about 30% of pediatric disease.^11,12^ Reviews of adult and pediatric cohorts report roughly 200 diagnostic genes overall, with a highly skewed distribution of yield: in early-onset CKD, seven genes account for about two thirds of diagnoses, and in adults, 10 genes accounting for more than 50% of the diagnostic yield.^13–16^

These findings have catalyzed the development of dedicated renal genetics clinics as an effective model to enhance the diagnostic process and clinical management of genetic kidney diseases. Now, specialized renal genetic clinics using next-generation sequencing (NGS) strategies and careful patient selection observe diagnostic yields of 20-40% in adult cohorts^11,12,14,17,18^ and as high as 65–67% for early-onset CKD, with even higher yields observed in cystic kidney diseases (77%) and tubulopathies (86%) compared to other disease categories^13,19–22^. These models outperform traditional workflows and highlight the critical role of stringent evidence-based inclusion criteria. The diagnostic yield is strongly determined by the composition of the cohort, defined as the pretest enrichment. The key determinants include the proportion with a positive family history or consanguinity, age at onset, the kidney disease suspected diagnosis at referral, CKD stage and severity, and the presence of extra-renal manifestations.^12,23,24^

These results informed the recent KDIGO guidelines (Kidney Disease: Improving Global Outcomes), which recommend routine genetic testing when CKD etiology is uncertain or when management may be altered by a genetic diagnosis.^25^ Importantly, economic evaluations in kidney genetics and in pediatric suspected monogenic disease show that earlier use of comprehensive sequencing is cost-saving or cost-effective, particularly in early-onset presentations.^22,26^ It is important to note, however, that these cost-effectiveness analyses considered only confirmed diagnostic findings and did not account for the potential downstream implications of variants of uncertain significance (VUS), which may generate additional healthcare costs, increase patient anxiety, and prompt unnecessary or potentially harmful investigations. Importantly, the rate of reporting variants of VUS is not uniform across populations or genes. It is higher among individuals from ancestries underrepresented in population reference databases and in genes that have historically been sequenced less frequently, where limited prior evidence constrains confident variant interpretation.^27^ Broad testing strategies using large renal panels or exome-backbone virtual panels are now common; however, larger static panels are associated with higher VUS rates, whereas exome and genome sequencing typically yield fewer VUS per test owing to more selective reporting practices.^28^ To our knowledge, no published cost-effectiveness analysis has explicitly compared curated gene-panel strategies with exome-based approaches while also modelling the potential downstream consequences of VUS (e.g., further testing and healthcare costs) into the outcome. In this study, we present a retrospective analysis of adult and pediatric patients with kidney disease who were evaluated for genetic testing in the multidisciplinary Nephrogenetics Clinic at the McGill University Health Centre in Montreal, Canada. We also describe how post-test variant adjudication, family-based testing, and standardized handling of secondary findings contribute to a more integrated model of genetic care. Our findings highlight the role of multidisciplinary renal genetics clinics in the diagnosis and management of inherited kidney disease.

## Methods

### Renal Genetics Clinic

In July 2019, a Renal Genetics clinic was created within the Division of Medical Genetics at the McGill University Health Centre (MUHC). The MUHC is an academic tertiary and quaternary care centre in Montreal, Quebec, Canada, and serves as the primary referral centre for the Réseau universitaire intégré de santé McGill (RUIS McGill). The clinic’s catchment area encompasses approximately 1.8 million people across seven regional health authorities spanning 63% of Quebec’s territory, reflecting the geographic and demographic diversity of the province. Educational talks and meetings were established with the faculty and staff within the Division of Nephrology to optimize the workflow of the Renal Genetics clinic and to familiarize nephrologists on referral criteria and genetic topics commonly encountered in the clinic. Eligibility for referral reflected standard Nephrogenetics practice: any clinical suspicion of a heritable kidney disorder, including early-onset CKD or ESKD at age 50 years or younger, positive family history, congenital anomalies of the kidney and urinary tract, cystic kidney disease, persistent hematuria or proteinuria, steroid-resistant nephrotic syndrome or focal segmental glomerulosclerosis, syndromic or extrarenal features, and unexplained tubulopathies or recurrent nephrolithiasis, and nephropathy of unknown etiology.^29^

Within the Renal Genetics Clinic, patients were evaluated by a medical geneticist or a genetic counsellor. Initial consultations included obtaining a comprehensive medical and three-generation family history, performing a personalized genetic risk assessment, and when appropriate, conducting a focused physical examination. Informed consent was obtained prior to testing, and sample collection (via blood or saliva) was coordinated for clinical genetic testing when indicated. Referrals were accepted from internal MUHC, external healthcare providers, and self-referrals directly from patients (Figure 1).

**Figure 1.**
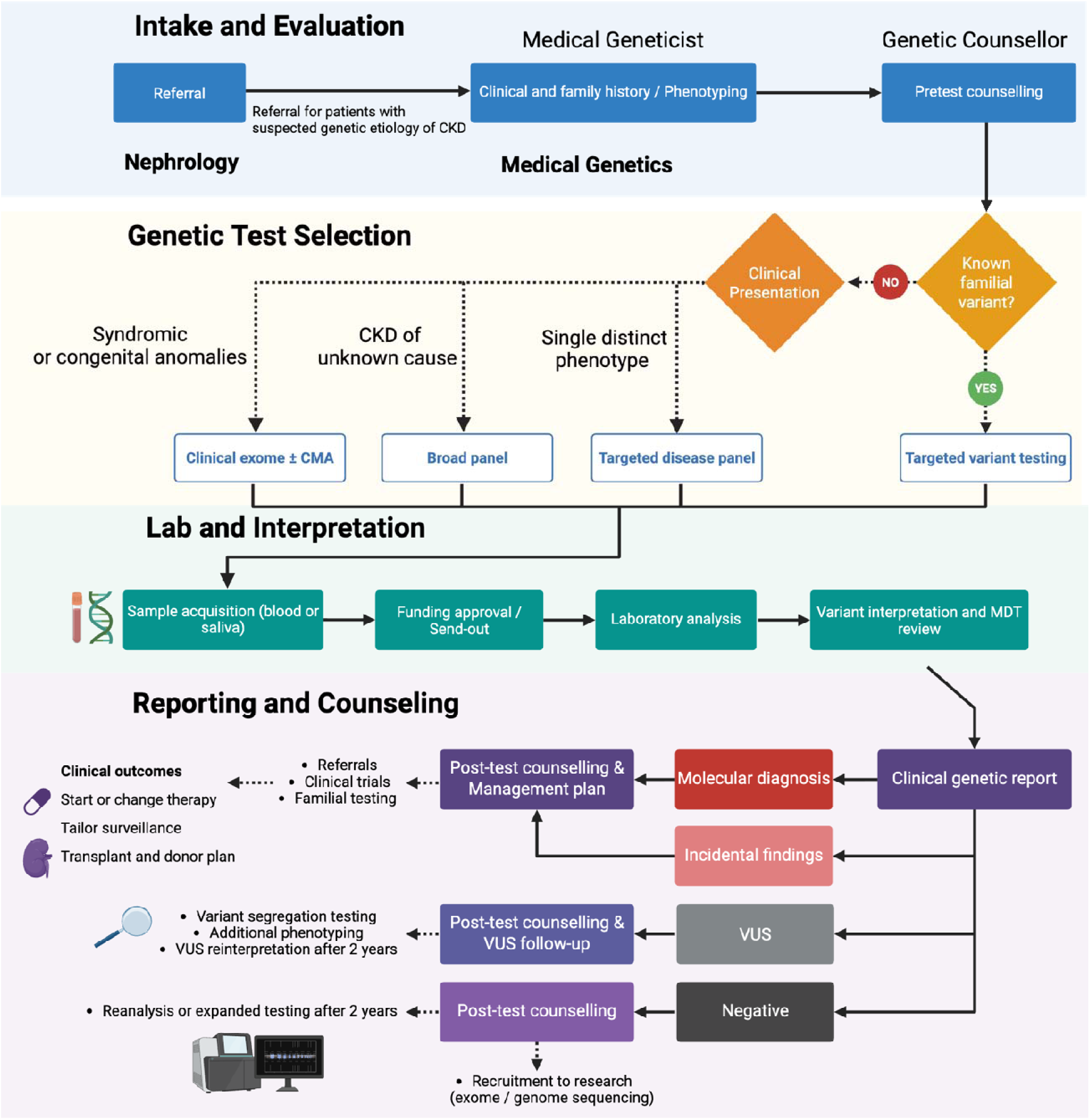
Renal genetics clinic workflow. The diagram shows referral and triage, medical genetics assessment and consent, phenotype-guided test selection, and sample collection with funding approval, followed by result-driven follow-up. Outcome pathways are depicted for positive, negative, variant of uncertain significance, and secondary findings, with corresponding actions including management updates, cascade testing, segregation studies, planned reinterpretation or reanalysis at 2 to 3 years, and research referral.

### Study Cohort

The present retrospective study included pediatric and adult patients with suspected inherited kidney disease, as defined by the referral criteria described above, who were evaluated in the Renal Genetics clinic and underwent clinical genetic testing between July 2019 and December 2024. We performed a retrospective chart review to collect data on demographic variables (age, self-reported race, and sex assigned at birth), clinical characteristics, detailed three-generation family history, referral indications, genetic testing results and implications for clinical management. The *a priori* clinical diagnosis was established based on broad etiologic disease categories, as determined by the referring nephrologist or by a medical geneticist with expertise in nephrology.

Clinical utility was assessed across five predefined categories: 1) tr*eatment modification*, defined as initiation or adjustment of therapy attributable to genetic diagnosis; 2) *transplant management changes*, defined as alterations in peri-transplant evaluation, including donor selection and perioperative protocols; 3) *living donor risk assessment*, defined as genetic evaluation to inform donor eligibility and counseling; 4) *cascade testing*, defined as targeted testing of at-risk family members; and 5) *prognostic clarity*, defined as refinement of prognosis and reproductive risk counseling.

Ethics approval for the retrospective chart review was obtained from the McGill University Health Centre Research Ethics Board (Protocol 2025-10930). For participants enrolled for reanalysis and expanded testing, and for the presentation of case-specific clinical information, approval was obtained from the same board (Protocol 2023-9387). Written informed consent was obtained in accordance with the Declaration of Helsinki.

### Genetic Testing

All diagnostic genetic tests ordered through the Renal Genetics Clinic were performed in CLIA (Clinical Laboratory Improvement Amendments)-certified laboratories, and variants were classified according to the respective standardized laboratory protocols. Most tests were ordered from Prevention Genetics (Marshfield, Wisconsin, United States) and Blueprint Genetics (Helsinki, Finland) as multi-gene panels. Blood or saliva swabs were collected for genetic testing at the time of in-person visit. For patients seen remotely via telemedicine, saliva collecting kits were shipped to their home.

Genetic tests ordered included targeted disease panel (<100 genes), broad multi-gene panels (>100 genes), phenotype-driven clinical exome-wide sequencing (virtual panel), chromosomal microarrays, single-gene sequencing and targeted variant testing. Quebec has a publicly funded health care system which covers the cost of gene panel testing for eligible patients, ranging from single-gene panels to phenotype-driven panels that include all OMIM-morbid genes.

### Genetic Testing Results

A positive diagnostic result was defined as the presence of a pathogenic or likely pathogenic (P/LP) variant in a gene that either fully or partially accounts for the patient’s clinical features. A positive nondiagnostic result referred to P/LP variants that were not causative of the patient’s kidney disease. An incidental finding was characterized by a P/LP variant in a gene associated with a condition unrelated to the individual’s kidney disease.

Specific genetic findings were classified as positive diagnostic results: heterozygous P/LP variants in *COL4A3* or *COL4A4*, as well as heterozygous *COL4A5* variants in females. *APOL1* high-risk genotypes (G1/G1, G1/G2, G2/G2) were counted as an etiologic diagnosis only when the clinicopathologic picture was concordant with *APOL1*-associated kidney disease, for example primary focal segmental glomerulosclerosis or collapsing glomerulopathy, and only after excluding an alternative monogenic or secondary cause. We documented such cases as “*APOL1*-associated nephropathy” rather than a Mendelian diagnosis, and in all other contexts recorded the genotype as a risk factor only. Cascade genetic testing was offered to all families with a positive result.

In cases with a positive diagnostic genetic result, two independent genetics professionals—a board-certified medical geneticist and a certified genetic counsellor—reviewed the initial clinical diagnosis in light of the genetic findings. Each case was then categorized into one of four groups: (1) *confirm*, where the genetic result aligned with the initial clinical impression; (2) *diagnose*, where a genetic cause was identified in patients without a known clinical diagnosis; (3) *reclassify*, where the genetic result led to a change in the presumed diagnosis; and (4) *at risk*, where the genetic finding indicated a predisposition to features not yet observed clinically.^30^

### VUS Stratification

All reported variants of uncertain significance were subclassified into VUS-low, VUS-mid, or VUS-high tiers using the evidence-level framework proposed by Rehm *et al.,* with each VUS scored accordingly to the points-based system proposed by Tavtigian *et al.* ^31^

### Total Points Classification

|  |  |
| --- | --- |
| ≥10 | Pathogenic |
| 6–9 | Likely pathogenic |
| 4-5 | VUS-high |
| 2-3 | VUS-mid |
| 0-1 | VUS-low |
| ≤ -1 | Likely benign |
| ≤ -7 | Benign |

## Statistical Analysis

Categorical variables were summarized as counts and percentages, and continuous variables as median with interquartile range. Diagnostic yield was defined as the proportion of probands with a pathogenic or likely pathogenic variant concordant with the clinical phenotype. Comparisons of diagnostic yield across referral categories, testing modalities, and demographic subgroups used Fisher’s exact test. All tests were two sided with *P* < 0.05 considered statistically significant. Analyses were performed in R version 4.4.0.

## Results

### Clinical Characteristics

A total of 206 probands with suspected hereditary kidney disease were included in this retrospective review. Patients were primarily referred from nephrologists with a clinical suspicion of an underlying genetic cause for their kidney disease. The cohort consisted of 170 adults (≥18 years) and 36 pediatric patients (<18 years). The median age at CKD diagnosis was 30 years (IQR: 18-45). The study population was ethnically diverse, reflecting the multicultural composition of the urban Montreal catchment area (Table 1). CKD of unknown etiology (CKDu) was the most common clinical presentation (58, 28%), followed by cystic nephropathy (52, 25%) and glomerulopathy (45, 22%). A family history of kidney disease was reported in 70% of patients, and extrarenal manifestations were present in 67%. At the time of referral, most patients were in an early stage of CKD (29% with mild-moderate (CKD 1-2), 16% with moderate-severe (CKD 3), 26% with severe-end-stage (CKD 4-5), and 5% post-transplant.

**Table 1.**
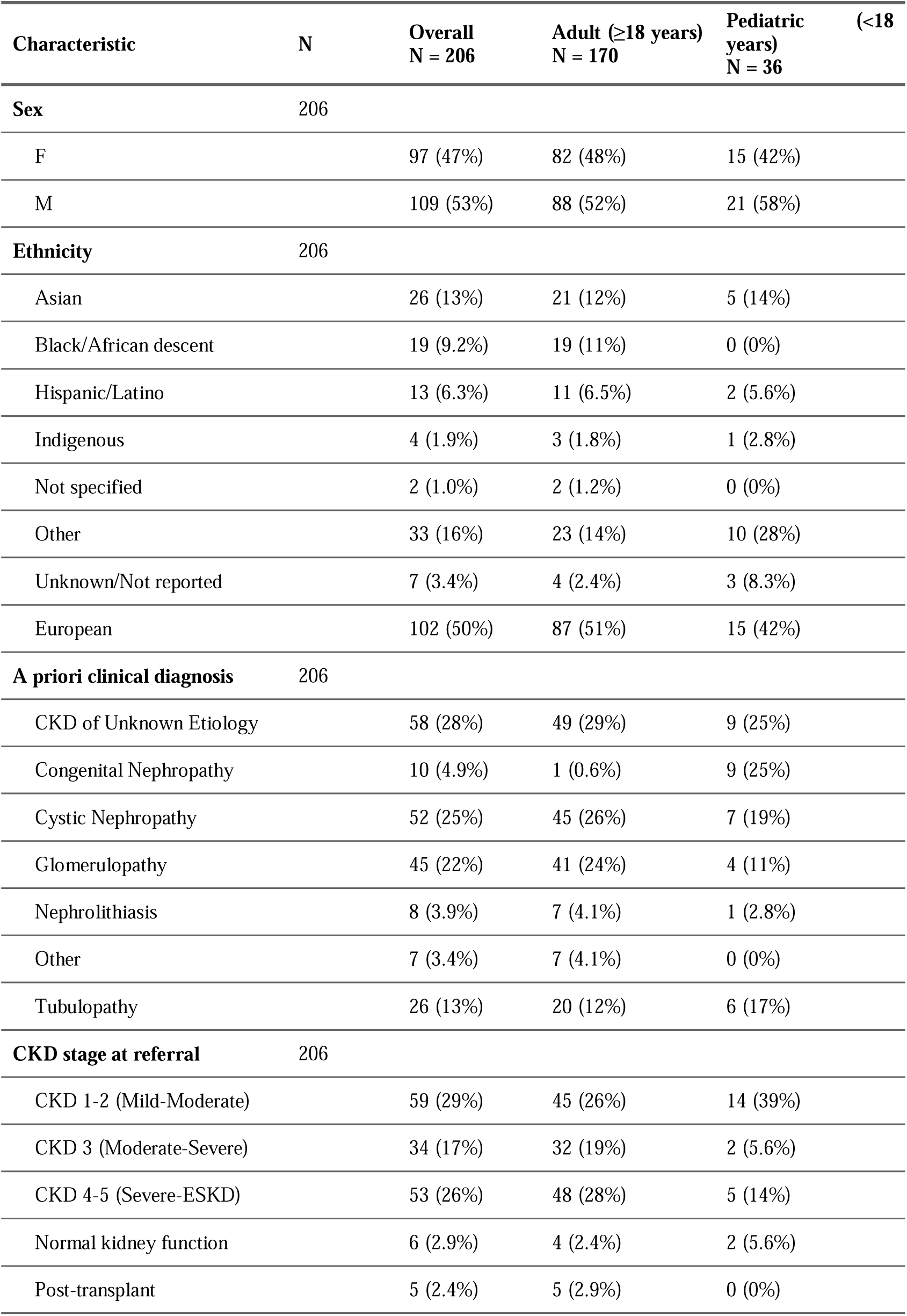
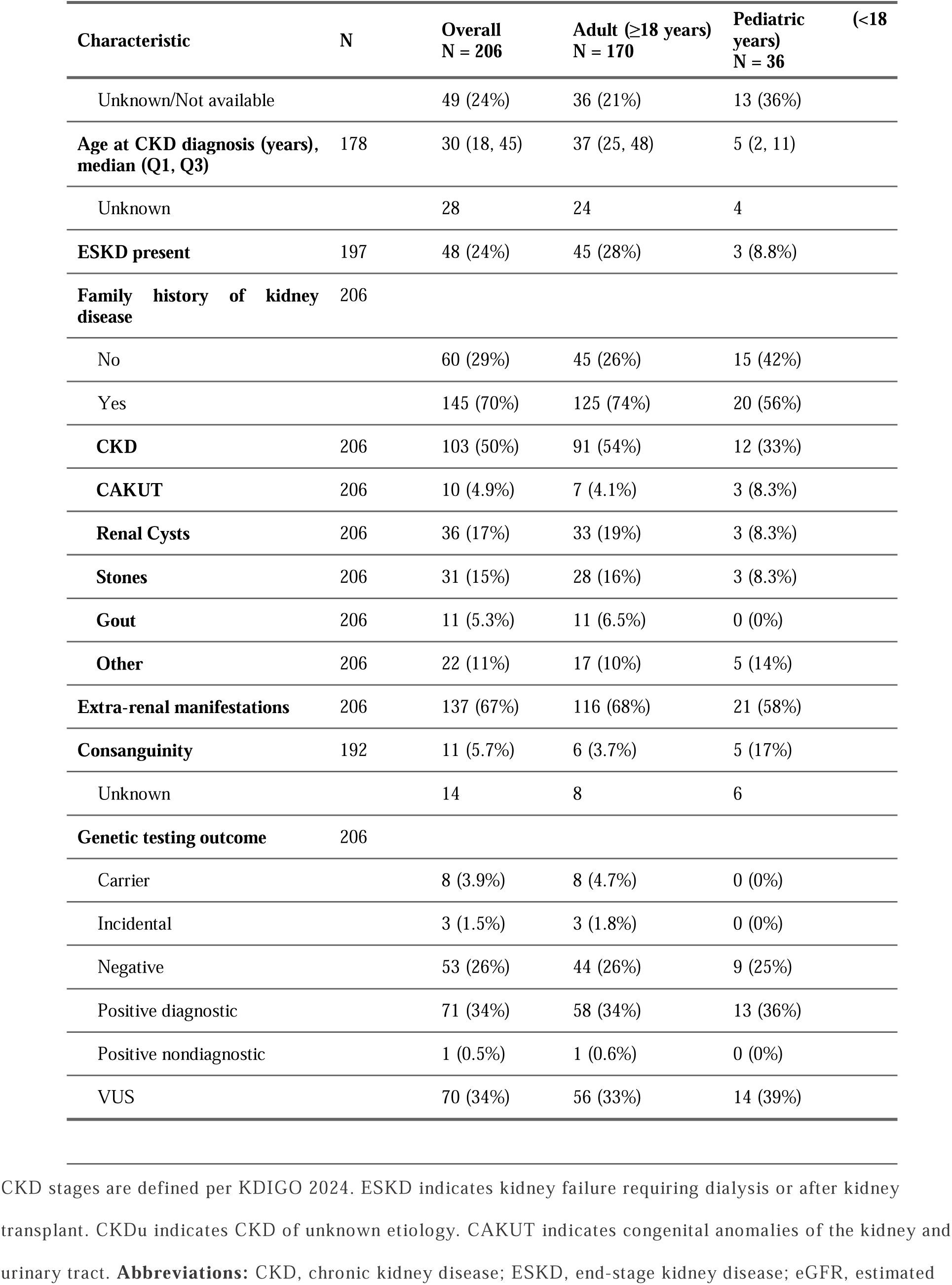
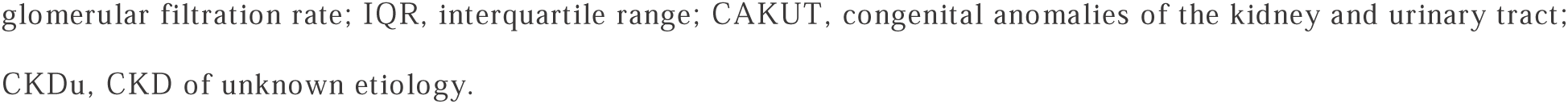
Clinical and Demographic Characteristics of Probands.

### Diagnostic Yield of Genetic Testing

Across 206 probands, 71 received a positive diagnostic result (34.5%), 8 were carriers only (3.9%), and 3 had incidental actionable findings (1.5%), and 1 had a positive non-diagnostic finding, for a total of 83 probands with any positive finding (40.3%) spanning 35 disease genes (Figure 2). The most frequently identified genes included *PKD1* (13 cases, 18.23%), *COL4A4* (6 cases, 8.8%), *HNF1B* (6 cases, 8.8%), and *PKD2* (5 cases, 7.4%) (Table 2). The distribution of diagnostic genes varied according to the age at CKD diagnosis, with *PKD1* and *PKD2* (autosomal dominant polycystic kidney disease) and heterozygous *COL4A3*, *COL4A4*, and *COL4A5* variants (autosomal dominant and X-linked Alport spectrum) predominating among adult-onset cases (Figure 2F).

**Figure 2.**
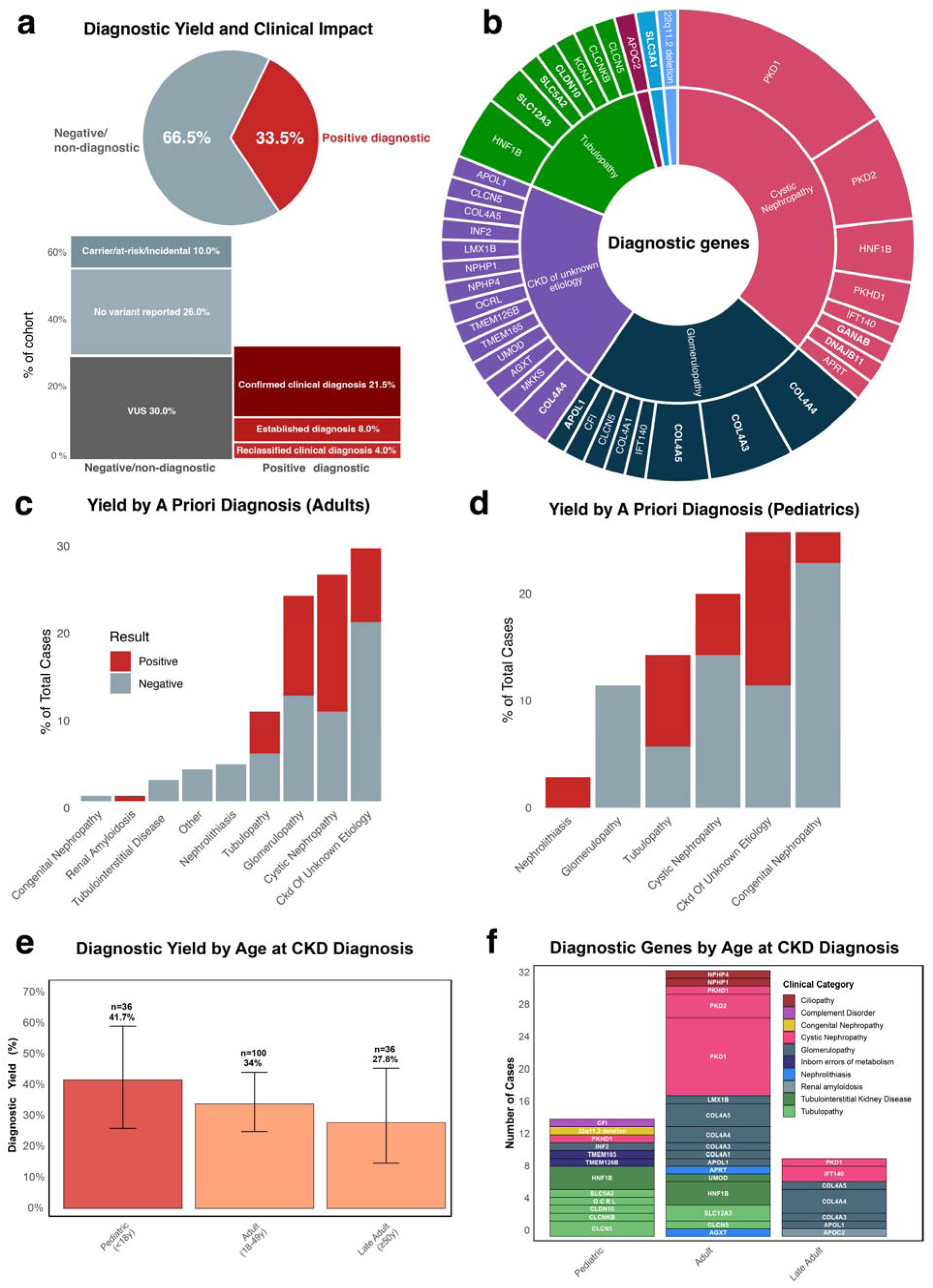
Diagnostic yield of genetic testing in patients with suspected hereditary kidney diseases. **A.** Pie chart showing overall diagnostic yield. Stacked bar chart below provides detailed subcategorization of results showing specific clinical outcomes including confirmed diagnoses, established diagnoses, reclassified diagnoses, partial diagnoses, identified carriers/at-risk/incidental findings, variants of uncertain significance (VUS), and cases with no variant reported. Numbers and percentages are shown for each category. **B.** Sunburst plot representing the diagnostic genes by proportion within each major disease category. Genes in bold are found in multiple disease categories. **C-D.** Diagnostic yield by a priori clinical diagnosis in adult patients (left) and pediatric patients (right). Stacked bar chart shows the proportion of positive (light blue) versus negative (light red) genetic testing results for each pre-test clinical diagnosis category in each cohort. **E.** Bar chart showing diagnostic yield by age at CKD diagnosis, with pediatric onset (<18 years) showing highest yield (41.7%). **F.** Stacked bar chart showing diagnostic genes categorized by disease category for each age-group. Error bars represent 95% confidence intervals. Numbers above bars indicate sample size and diagnostic yield percentage. Data shown for probands with CKD stage data available. Diagnostic yield was high overall and enriched in cystic, tubulopathy, and glomerulopathy referrals, with pediatric-onset cases showing the highest yield and adult diagnoses dominated by ADPKD and *COL4A*-related Alport spectrum, reflecting referral enrichment and disease architecture.

**Table 2.**
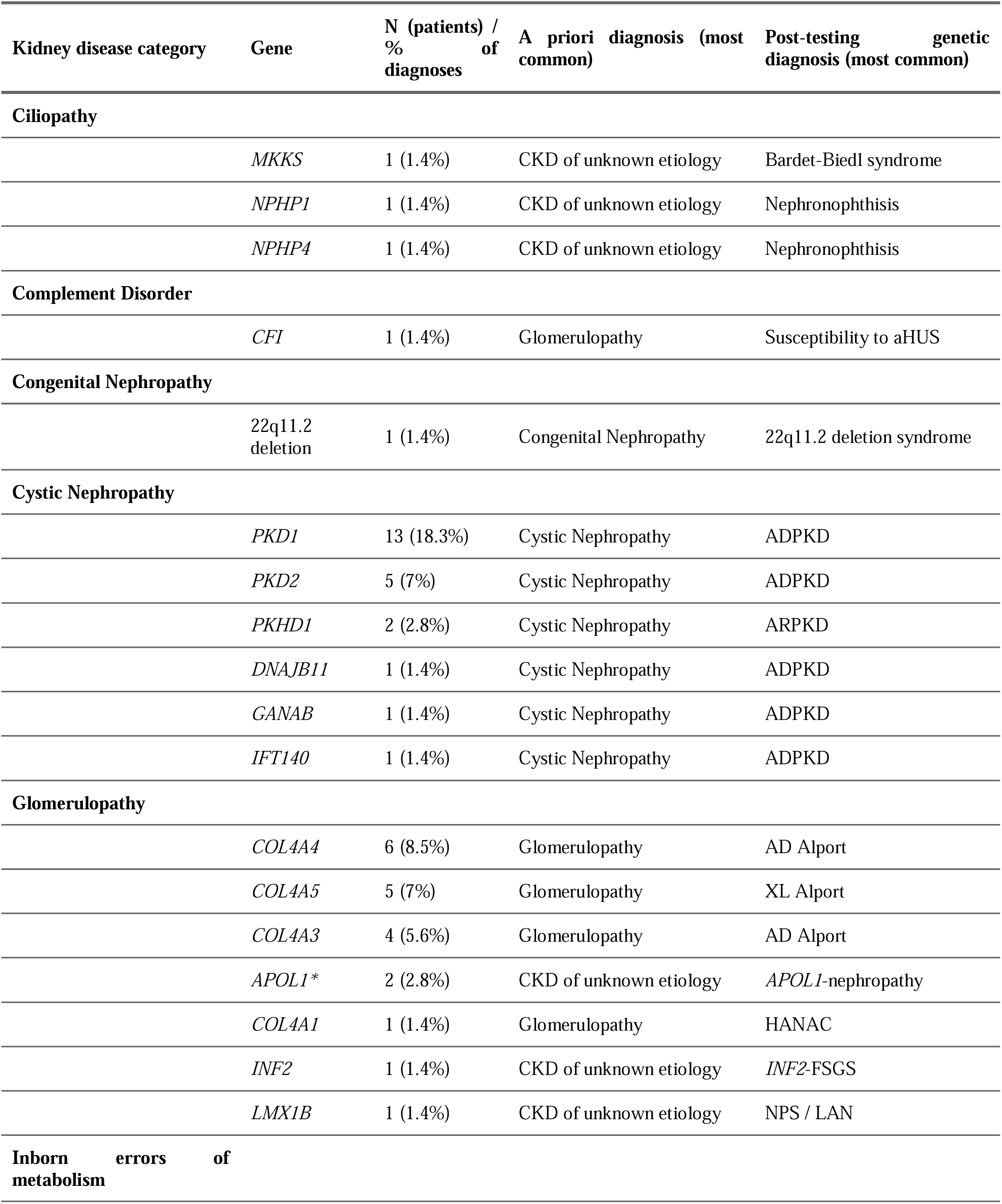
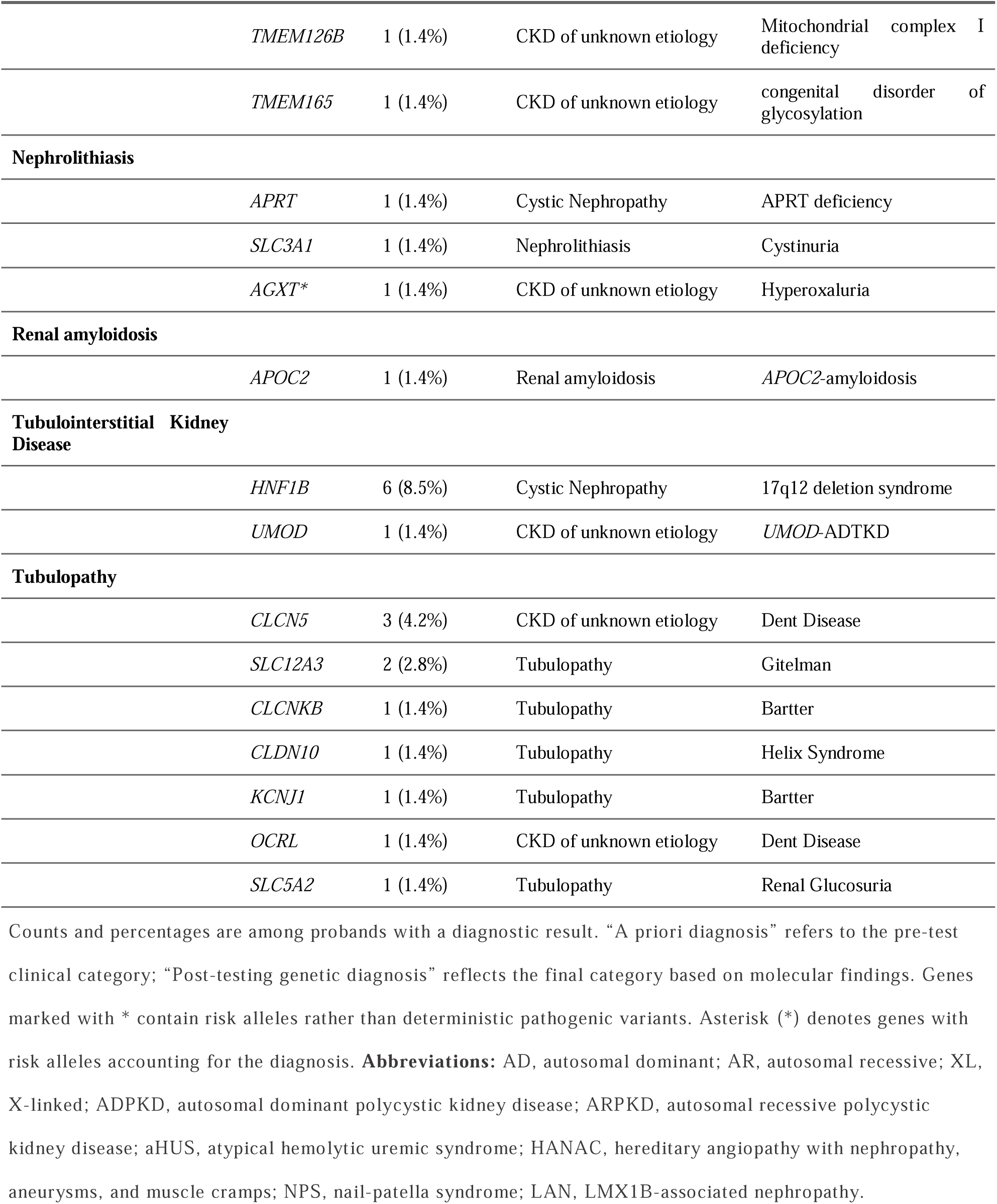
Summary of Diagnostic Genes.

The highest diagnostic yields were observed in cystic nephropathy (51.9%), tubulopathy (38.5%) and glomerulopathy (35.6%) (Table 3). Diagnostic yield was 42% in cases with pediatric CKD onset, 34% in younger adults (18-49 years), and 27.8% in older adults (≥49 years) at CKD onset; with no statistically significant difference in diagnostic yield across age groups (Fisher’s exact test). A total of 97 VUS were identified in 81 probands (39%), most frequently in *PKD1* (n=10).

**Table 3.** Clinical Outcomes of a Diagnosis.

| Clinical diagnosis | N | Diagnosis<br>impacting clinical<br>care | Change in<br>management | Changed<br>treatment<br>plan | Informs<br>risk for<br>biological<br>donation | Change in<br>kidney<br>transplant<br>management | Cascade<br>testing in<br>family | Early<br>diagnosis for<br>family<br>member | Provides<br>prognostic<br>clarity |
| --- | --- | --- | --- | --- | --- | --- | --- | --- | --- |
| Cystic<br>Nephropathy | 25 | 25 (92.6%) | 4 (14.8%) | 2 (7.4%) | 5 (18.5%) | 2 (7.4%) | 22 (81.5%) | 1 (3.7%) | 27 (100%) |
| CKDu | 15 | 14 (87.5%) | 5 (31.2%) | 2 (12.5%) | 1 (6.2%) | 0 (0%) | 9 (56.2%) | 3 (18.8%) | 16 (100%) |
| Glomerulopathy | 15 | 13 (86.7%) | 5 (33.3%) | 4 (26.7%) | 4 (26.7%) | 2 (13.3%) | 9 (60%) | 1 (6.7%) | 13 (86.7%) |
| Tubulopathy | 10 | 10 (100%) | 2 (20%) | 3 (30%) | 0 (0%) | 0 (0%) | 4 (40%) | 2 (20%) | 10 (100%) |
| Congenital<br>Nephropathy | 1 | 1 (100%) | 0 (0%) | 0 (0%) | 0 (0%) | 0 (0%) | 1 (100%) | 0 (0%) | 1 (100%) |
| Nephrolithiasis | 1 | 1 (100%) | 1 (100%) | 1 (100%) | 0 (0%) | 0 (0%) | 1 (100%) | 0 (0%) | 1 (100%) |
| Renal<br>amyloidosis | 1 | 1 (100%) | 0 (0%) | 0 (0%) | 0 (0%) | 0 (0%) | 1 (100%) | 0 (0%) | 1 (100%) |
| TOTAL | 68 | 65 (91.5%) | 17 (23.9%) | 12 (16.9%) | 10 (14.1%) | 4 (5.6%) | 47 (66.2%) | 7 (9.9%) | 69 (97.2%) |

Summary of clinical outcomes for patients with positive diagnostic genetic results, stratified by clinical diagnosis category. "Change in management" represents any clinical action resulting from the genetic diagnosis, while "Changed treatment plan" refers to initiation or modification of therapeutic interventions (subset of management changes). Additional management changes include transplant planning adjustments. Numbers and percentages shown for each outcome category. **Abbreviations:** CKDu, CKD of unknown etiology.

### Clinical Outcomes of Genetic Testing

Genetic testing played a pivotal role in resolving or reclassifying the clinical diagnosis in several cases. Among patients initially categorized as CKDu (n=58), genetic testing identified a definitive molecular diagnosis in 25.9% of cases, most often within the glomerulopathy subgroup (Figure 3).

**Figure 3.**
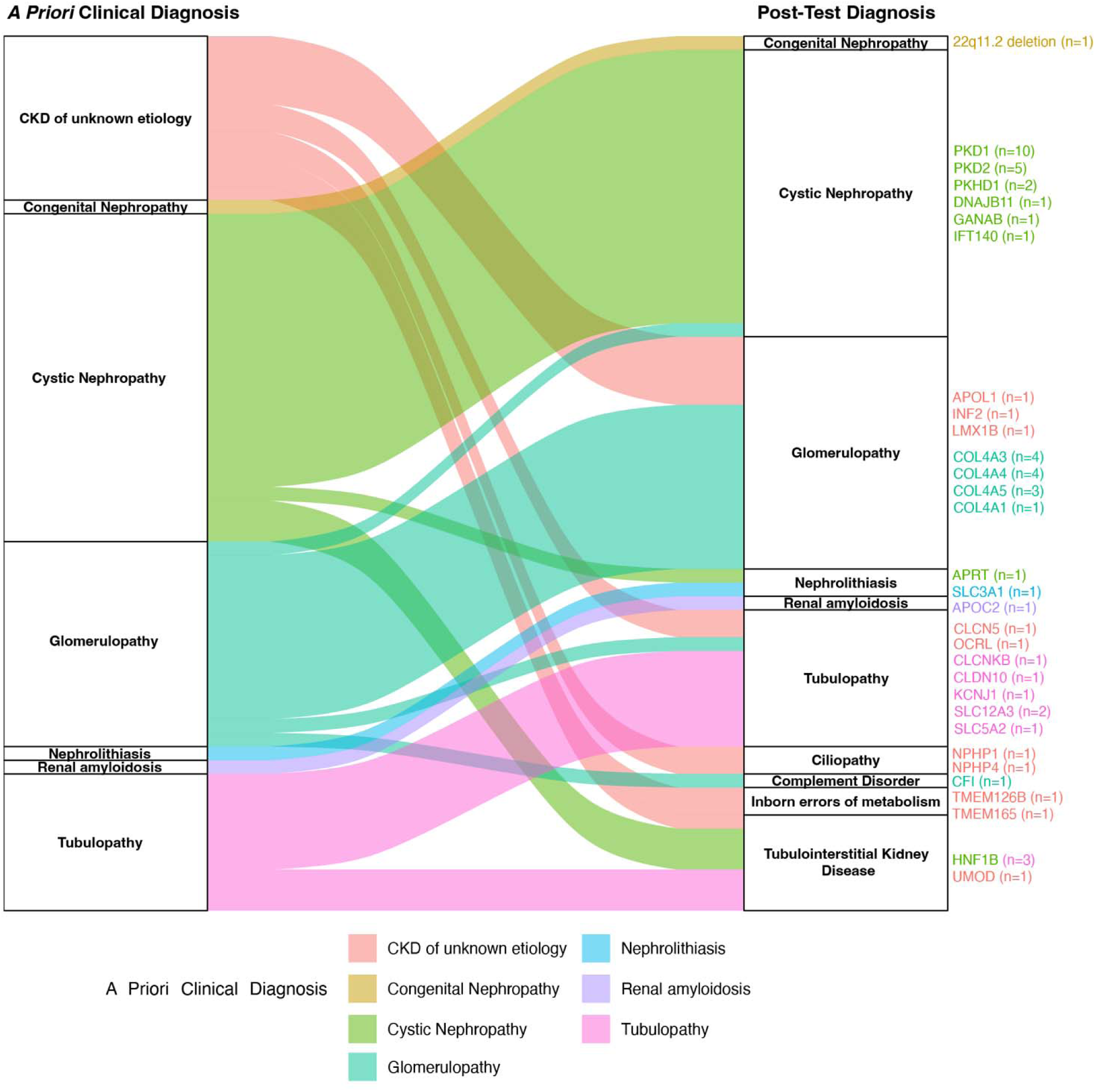
Diagnostic Reclassification: Pre-test (*A Priori*) vs Post-test (*Post hoc*) Diagnosis. Alluvial plot illustrating diagnostic reclassification flow from a priori clinical diagnosis (left) to final kidney disease category classification (right) for patients with positive diagnostic genetic testing results. Each flow stream represents patients, with stream thickness proportional to the number of cases. Colors correspond to the original a priori clinical diagnosis. On the right, the diagnostic genes associated with each clinical category are shown with the number of likely pathogenic/pathogenic variants represented in parentheses. Genetic testing frequently converted nonspecific labels, especially CKD of unknown etiology, into specific molecular entities, thereby resolving diagnostic uncertainty and aligning care with gene-defined pathways.

Across the entire cohort, three unrelated individuals were found to have incidental but medical actionable genetic findings that redirected non-renal management. One illustrative case involved an adolescent transplant candidate with end-stage kidney disease, peripheral neuropathy, and ventricular trabeculations on echocardiography. Phenotype-guided exome-wide panel sequencing identified biallelic pathogenic variants in *PKP2*, establishing a predisposition to arrhythmogenic right ventricular cardiomyopathy (OMIM #609040), a hereditary cardiac disorder predisposing to ventricular arrhythmia and sudden cardiac death. Cardiology referral, implementation of exercise restriction, and cascade testing for at-risk relatives were initiated, and peri-transplant cardiac monitoring was incorporated into his care plan.

Genetic results directly influenced clinical management in 65 of 71 molecularly diagnosed probands (91.5%) (Table 3). The most frequent interventions included treatment modification (16.9%), such as initiation or confirmation of renin–angiotensin system blockade in Alport syndrome, commencement of xanthine oxidoreductase inhibitors following confirmation of Adenine Phosphoribosyl Transferase Deficiency, and use of mitochondrial-targeted supplements following molecular diagnosis of mitochondrial disorder. Additional management changes involved evaluation of living-donor risk (14.1%), and revision of transplant planning, including adjustment of anesthesia protocols (5.6%). Cascade testing was initiated in 44 families (66.2%) and identified at-risk relatives in seven (9.9%). All solved cases received detailed prognostic and reproductive risk counselling.

A representative clinical vignette highlights the broader impact on families. A male (early 30s) with persistent microhematuria underwent targeted sequencing of *COL4A3, COL4A4*, and *COL4A5,* which revealed a heterozygous likely pathogenic *COL4A3* variant and confirmed autosomal-dominant Alport syndrome (historically referred to as thin basement membrane nephropathy). Post-test counselling prompted baseline audiology and retinal assessment and a plan for supportive management, with initiation of renin angiotensin system blockade if albuminuria develops, consistent with current clinical guidelines.^32^ Although the lifetime risk of kidney failure for heterozygous *COL4A3* or *COL4A4* carriers is generally considered low, these individuals remain at increased risk of proteinuria, hypertension, and reduced kidney function, particularly when there is a positive family history of CKD. Accordingly, life-long renal surveillance and detailed family assessment were arranged. Carrier screening in the patient’s partner identified a different *COL4A3* pathogenic variant, prompting her own annual monitoring for proteinuria and a discussion of reproductive options, such as *in vitro* fertilization with pre-implantation genetic testing. During a subsequent pregnancy, amniocentesis demonstrated that the fetus had inherited the paternal variant, thereby likely reducing the risk of autosomal-recessive Alport syndrome and permitting routine obstetric management. This case underscores the role of genomics in resolving diagnostic uncertainty and enabling precision management in nephrology.

### Variants of Uncertain Significance: A Significant Diagnostic Challenge

Minority ancestry groups exhibited lower diagnostic yields and a higher mean burden of VUS per individual (Figure 4A–B), consistent with their underrepresentation in population databases and the reduced applicability of the PM2 allele-frequency criterion. Overall, 39% of probands carried at least one VUS. Post-test internal reclassification of the reported VUSs was performed using the 7-point system proposed by Tavtigian *et al.* and the VUS tiering framework (VUS-low, VUS-mid, VUS-high) (Figure 4C).^28,31,33^ Of these, 61 of 90 (67%) were categorized as VUS-mid or lower. Many of these variants showed limited genotype-phenotype concordance and are candidates for reclassification to *Likely Benign* as additional population or functional data emerge. Variants classified as VUS-high (n=21, 23.3%) demonstrated strong genotype-phenotype concordance but lacked sufficient evidence to meet pathogenicity thresholds, necessitating segregation or functional studies for confirmation. Several variants had accumulated enough supporting evidence to be reclassified as *Likely Pathogenic* or *Pathogenic* (n=8, 8.9%), underscoring the value of multidisciplinary variant review (Figure 4C). Notably, compound heterozygous *SLC12A3* variants (c.1967C>T p.Pro656Leu; c.1837G>A p.Gly613Ser) in a patient with hypokalemia and a clinical suspicion of Gitelman syndrome, met criteria for *likely pathogenic* upon re-evaluation.

**Figure 4.**
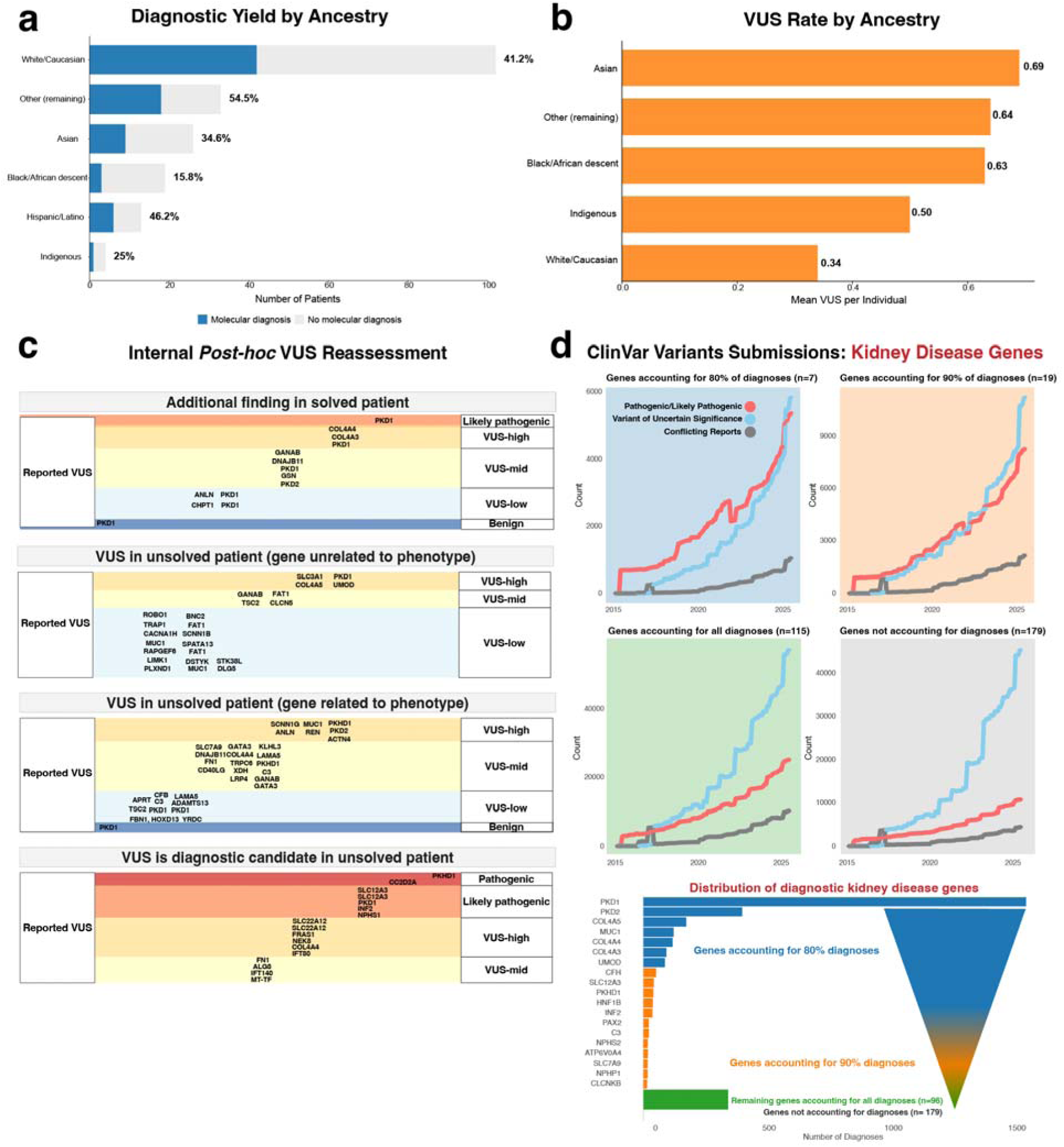
VUS and Diagnostic Analysis. **A.** Bar chart showing diagnostic yield by ancestry. Dark blue bars represent individuals with a molecular diagnosis (Partial Diagnosis, Confirmed Diagnosis, Reclassified Diagnosis, Candidate diagnosis, at-risk, Diagnosis) and dark red bars represent no diagnosis. Percentages and counts are displayed for each ancestry group, with a legend indicating outcomes. **B.** Bar chart depicting the mean number of variants of uncertain significance (VUS) per individual by ancestry group. **C.** Internal *post hoc* VUS reassessment applying ACMG/AMP with point-based scoring and evidence-level VUS subclassification in clinical context. **D.** ClinVar analysis of submitted Pathogenic/Likely pathogenic variants, Variants of uncertain significance, or Variants with Conflicting reports over time. Facets represent different categories of diagnostic renal disease genes reported by the meta-analysis from Schott *et al.*^14^, grouped by their cumulative diagnostic contribution to diagnosed patients. Individuals from underrepresented ancestries had lower diagnostic yield and higher VUS burden, and most reported VUS were mid or lower confidence on internal review, while high-confidence VUS clustered in well-established renal genes, underscoring the need for structured re-evaluation and ancestry-inclusive data generation.

### Inclusion of low-yield genes increases VUS burden

Large multi-gene panels generate VUS more frequently than exome or genome sequencing, and the VUS rate rises with panel breadth.^28^ To quantify this effect, we stratified kidney-disease genes by cumulative diagnostic yield, based an adult CKD meta-analysis, and examined ClinVar submission trajectories within each yield stratum.^14,34^ Figure 4D shows that high-yield genes accrued proportionally more pathogenic or likely pathogenic submissions relative to VUS, whereas lower-yield genes were associated with a steeper rise in VUS entries. This pattern indicates that inclusion of genes with limited evidence for disease causality contributes disproportionately to the overall VUS burden.

### Comparing the Clinical Validity of Genetic-Testing Strategies

#### Observed diagnostic yield in our cohort

Multi-gene panel testing represented the predominant diagnostic approach in our cohort. Patients without a strong suspicion of a specific disease etiology (e.g., CKDu) or those with negative results from targeted panels underwent either broad multi-gene panel testing encompassing all kidney disease genes or exome-based testing of clinically-indicated genes (Figure 5A). Among patients who underwent targeted disease panels as their first or only test (65/206), the diagnostic yield was 41.3%. Fourteen of these individuals underwent additional testing (e.g., broad testing), but only two additional diagnoses were obtained. In contrast, 54% (111/206) of individuals were initially tested using broad (comprehensive) kidney-disease panel first, which achieved a 28.9% diagnostic yield.

**Figure 5.**
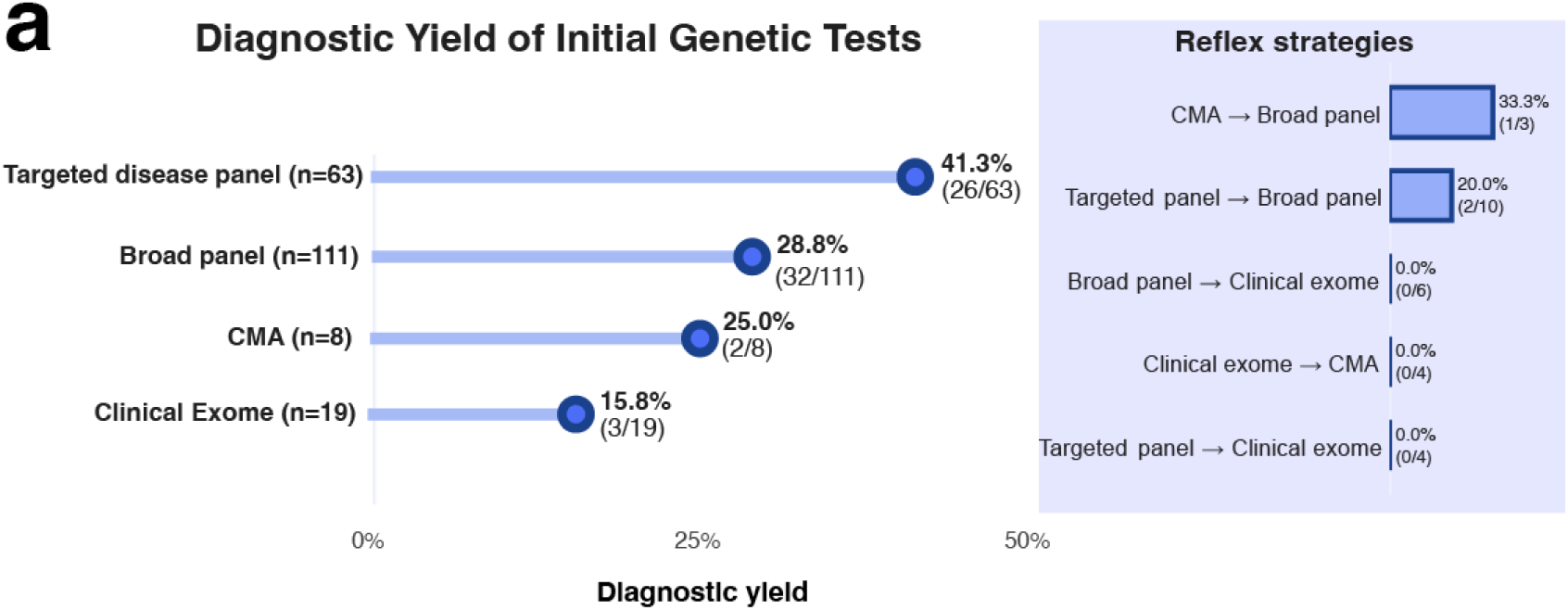
Diagnostic Pathways of Genetic Testing Strategies in Kidney Disease. **A.** Observed diagnostic pathways among probands, showing the sequence of testing and diagnostic outcomes for clinically ordered tests. *Inset:* Reflex strategies for probands receiving more than one genetic test. Arrow heads represent where a subsequent test was performed due to a negative result. Only pathways with ≥ 2 patients are displayed.

## Discussion

Genetic testing has emerged as an essential component of nephrology practice, yet the optimal implementation strategy and its real-world diagnostic and clinical impact remain incompletely defined. In this study, we evaluated the diagnostic and clinical impact of integrating a dedicated renal genetics service within a tertiary nephrology program, examining molecular yield, management outcomes, ancestry-linked disparities, and the comparative performance of different genetic-testing approaches. We observed a high molecular diagnostic rate of 34.5% with clear clinical utility. Among diagnosed probands, any management impact occurred in 91.5%, with treatment modifications in 16.9%, living-donor risk assessment in 14.1%, and peri-transplant adjustments in 5.6%. Cascade testing was initiated in 66.2% of families, extending benefits to relatives. Our single-centre experience also quantifies ancestry-linked disparities in diagnostic yield and VUS burden and compares testing strategies to guide implementation in diverse populations. We enriched referrals using multiple criteria, including early-onset CKD or kidney failure at ≤30 years, yet in adults our diagnostic yield did not vary by age at CKD onset. Clinical programs often use broader onset cutoffs such as <50 years, and recent meta-analytic data show substantial yield at ≥50 years and no independent association between younger age and yield, indicating that age alone should not restrict testing.^14,35–37^

The primary value of genetic testing in nephrology is to provide diagnostic certainty and guide actionable clinical decisions. This was most evident in patients with CKD of unknown etiology (CKDu), who comprised 28% of our cohort and for whom a definitive molecular cause was identified in 25.9% of cases, converting nonspecific labels to entities with defined prognostic and therapeutic pathways, consistent with other specialized programs.^30^ A genetic diagnosis informs treatment by confirming the need for therapies like renin-angiotensin system blockade in Alport syndrome or by preventing the use of unnecessary immunosuppression. In the context of transplantation, genetic results are critical for evaluating the eligibility of living-related donors and for tailoring peri-transplant care.^10^

The spectrum of *HNF1B*-related disease in our cohort illustrates the benefit of comprehensive analysis for tubulointerstitial disease and cystic presentations, including frequent 17q12 deletions and diverse extra-renal features, which are often under-recognized in adult nephrology.^38,39^ Six individuals were diagnosed with *HNF1B*-associated disease, five harboring the recurrent 17q12 microdeletion and one with a de novo frameshift variant (c.262_283dup p.Leu95Hisfs*29) presenting in infancy. Clinical phenotypes ranged from isolated magnesium-wasting tubulopathy to predominant cystic nephropathy, most accompanied by hypomagnesemia. Extrarenal manifestations were common and included pancreatic insufficiency, Müllerian duct anomalies, diabetes, strabismus, and calcium pyrophosphate deposition disease. Age at presentation spanned infancy to late adulthood, reflecting the substantial variable expressivity and frequent under-recognition of this multisystem disorder in adult nephrology practice.

High-risk *APOL1* genotypes (G1/G1, G1/G2, G2/G2) increase kidney disease risk with incomplete penetrance under a recessive model and are recommended to be reported as risk alleles rather than deterministic pathogenic variants.^40,41^ In transplantation, donor *APOL1* high-risk genotypes are associated with shorter deceased-donor graft survival in adjusted analyses and small living-donor cohorts suggest greater post-donation eGFR decline and occasional kidney failure among high-risk donors.^42^ In line with KDIGO recommendations to offer *APOL1* testing with shared decision-making while acknowledging uncertain individual risk, we did not count *APOL1* high-risk genotypes toward Mendelian diagnostic yield and instead report them separately to guide counseling and donor assessment.^43^

Our study highlights the strengths of embedding renal genetic testing within a medical genetics service. In this shared-care pathway, nephrologists refer patients to a centralized genetics team, which has proven effective and scalable in mainstream nephrology settings.^23,44^ Such a model standardizes pre-test genetic counseling, effectively manages secondary findings, and maintains a formal reanalysis pathway for VUS; it also enables testing beyond the primary renal phenotype when warranted (e.g., syndromic features).^45^ This was illustrated by a case in our cohort where an incidental finding of pathogenic variants in *PKP2* led to a new diagnosis of arrhythmogenic right ventricular cardiomyopathy, prompting life-saving changes to the patient’s peri-transplant care plan.

Variants of uncertain significance (VUS) are common and should not trigger management change. Overall, 39% of probands had at least one VUS, and 34% had a VUS-only result with no diagnostic variant reported. This rate is high relative to other renal genetic cohorts and is expected in a diverse clinic population that predominantly use broad multigene panels, which are associated with higher VUS rates and greater uncertainty in underrepresented ancestries.^23,28,46^ Individuals of non-European ancestry had a lower diagnostic rate and a higher VUS burden, consistent with limited representation of diverse populations in reference databases.^46^ We applied a systematic *post hoc* re-evaluation of all VUS using the ACMG/AMP point-based framework and an a priori evidence-tiering schema.^28,31,33^ This structured reassessment revealed that while the majority of VUS (67.4%) had low-to-mid-level evidence and are candidates for future downgrading or omission from the clinical report, a significant minority (31.4%) were classified as high-level evidence or met criteria for likely pathogenic, prioritizing them for targeted segregation or functional studies (Figure 4C). This approach yielded concrete upgrades in several cases, highlighting a critical message for nephrology practice: VUS require a formal, multidisciplinary re-evaluation pathway from genomics specialists to manage uncertainty responsibly, filter out variants of low clinical relevance, and focus resources on those with a high probability of being pathogenic.

VUS disproportionately arise from genes with weaker clinical validity, while genes with strong validity more often contained pathogenic or likely pathogenic variants (Figure 4D). Furthermore, VUS categorized as VUS-mid or lower are enriched within these low-evidence genes that are included in comprehensive panels but are rarely or never reported as diagnostic (Figure 4C). Conversely, VUS classified into the VUS-high category, or with evidence for reclassification to likely pathogenic, are found in well-established disease-causing genes (e.g., *PKHD1, SLC12A3, PKD1*). This VUS burden can be prospectively mitigated by restricting panels to phenotype-matched, clinically validated genes or by utilizing an exome backbone with virtual panels that enable same-sample expansion after a negative analysis. This strategy follows ACMG and ClinGen guidance to limit clinical action to pathogenic or likely pathogenic variants and to manage VUS with structured follow-up, not immediate clinical intervention.^47–49^

### Limitations

This study represents a single-centre, referral-based renal genetics service comprising probands clinically referred for suspected monogenic kidney disease in an urban, multicultural setting. The referral-criteria enrichment increases the pretest probability of detecting a genetic diagnosis relative to unselected CKD populations and is therefore appropriate for assessing diagnostic yield and clinical utility within a genetics clinic context. The cohort does not reflect population-level prevalence or representativeness. Outcome ascertainment relies on retrospective chart abstraction. Analyses are descriptive and unadjusted and use available-case denominators. If missing data are not completely at random, the resulting estimates can be biased.

### Considerations for Implementing a Renal Genetics Clinic

This Canadian single-centre experience in a diverse population highlights the clinical value of integrating a specialized genetics service into nephrology care. Our findings demonstrate that a referral process based on defined clinical criteria is effective for achieving a high diagnostic yield and supporting the personalized management of hereditary kidney disease. Implementing a renal genetics clinic with a multidisciplinary genomic team can improve diagnostic accuracy and inform management in more than 90% of cases.^30^ This collaborative process integrates nephrology, genetics, and pathology expertise, facilitates *post hoc* phenotyping and the strategic use of familial segregation studies to reclassify variants and guide clinical management.^50^ This can reduce the need for subsequent analyses and patient visits, and this approach ensures that the clinic team is qualified to handle interpreting and communicating ambiguous findings.

Based on this, we make the following recommendations for implementing a multidisciplinary renal genetics practice:

1. **Build a synergistic shared nephrology-genetics pathway:** Nephrologists triage and refer to a genetics clinic. The genetics team performs deep phenotyping and pretest counseling, selects and orders the test, and liaises with the laboratory. Results are interpreted in a joint review by a medical geneticist and a genetic counselor with nephrologist input, returned with a documented management plan, and routed to relevant services. The genetics clinic coordinates cascade testing and transplant donor assessment when indicated.
2. **Criteria for considering genetic testing:**

a. CKD of unknown etiology after standard work-up.
b. Positive family history of CKD, dialysis, kidney transplant, or a known hereditary renal disorder, or a pedigree consistent with Mendelian disease.^25^
c. Early-onset CKD (≤ 50 years) or syndromic features including extra-renal manifestations or consanguinity.^25^
d. Persistent hematuria or steroid-resistant proteinuria with suspected Alport spectrum; offer cascade testing of first-degree relatives when *COL4A3, COL4A4,* or *COL4A5* variants are identified.^32,51^
e. Cystic kidney disease with uncertain diagnosis by imaging.^52^
f. Evaluation of at-risk living related donors.^53^
3. **Broad but deep first-tier testing strategy:** Define first-tier testing using a curated renal gene set limited to disorders with moderate or strong ClinGen gene–disease validity to minimize false positives and VUS burden.^54^ Testing must provide complete coverage of all included genes and known pathogenic variant classes, supplemented by targeted assays for technically challenging loci when necessary. In resource-limited, publicly funded systems, an exome backbone with phenotype-driven virtual panels offers an optimal compromise, enabling same-sample expansion after negative results while controlling VUS reporting and costs.^55^ Gene panels provide lower upfront costs and established coverage of technically challenging disease-causing genes (e.g., *PKD1, MUC1, CFHR1-5*), whereas exome-first sequencing facilitates same-sample expansion, trio analysis, and streamlined periodic reinterpretation, but may require augmented methods for duplicated regions and structural variants.
4. **Holistic genetic care, family management, variant interpretation and follow-up:** Adopt a genetics-led, family-centered model that integrates pre-test counseling with documentation of preferences for secondary findings, standardized ACMG secondary-findings management, and clear consent workflows.^45^ Centralize variant interpretation to apply ACMG/AMP rules, treat VUS as non-actionable, and align genetic testing strategies based on the clinical presentation. Implement scheduled reanalysis and recontact programs. Systematically offer cascade testing and defined referral pathways, informing donor selection and surveillance strategies in relative. This model reduces inappropriate management changes, increases diagnostic yield and allocation of health-care resources, and converts a proportion of negative, or VUS results to diagnoses over time, improving safety and clinical utility.

## Data Availability Statement

Additional data can be shared by contacting the corresponding author upon reasonable request.

## Data Availability

All non-identifiable participant data underlying the results are available from the corresponding author upon reasonable request

## Acknowledgements

This work was supported by a Fonds de recherche du Québec – Santé Junior 1 Clinician-Scientist Award to T.K. and the SickKids New Investigator Research Grant (NI21-1159). Z.S. received a Canadian Institutes of Health Research Doctoral Research Award. Dr. Sandal is supported by the Fonds de recherche du Québec – Santé Junior 1. The funders had no role in study design, data collection, analysis, interpretation, or manuscript preparation

## Author Contributions

Zachary T. Sentell and Felicia Russo contributed equally to this work. Thomas M. Kitzler, Zachary T. Sentell, and Felicia Russo conceptualized the study. Thomas M. Kitzler and Felicia Russo reviewed and interpreted the clinical genetic data. Zachary T. Sentell conducted the formal research analysis, developed the methodology, and drafted the original manuscript. Felicia Russo curated the data and contributed to the formal analysis and writing. Ruth Sapir-Pichhadze contributed to the economic analysis and results interpretation. Marc Henein, Zachary W. Nurcombe, Lina Mougharbel, and Elena Torban contributed to data interpretation and critically reviewed the manuscript. Ahsan Alam, Lorraine E. Bell, Daniel Blum, Dana Baran, Marcelo Cantarovich, Andrey V. Cybulsky, Sonali De Chickera, Mallory L. Downie, Bethany J. Foster, Gershon Frisch, Paul R. Goodyer, Indra R. Gupta, Laura Horowitz, Serge Lemay, Mark L. Lipman, Sharon J. Nessim, Tiina Podymow, Ratna Samanta, Shaifali Sandal, Ruth Sapir-Pichhadze, Rita Suri, Tomoko Takano, Emilie Trinh, and Murray Vasilevsky provided patient resources and clinical data from the renal genetics cohort. Thomas M. Kitzler provided supervision, critically revised the manuscript, and was responsible for funding acquisition. All authors reviewed and approved the final manuscript.

